# Spread of infection and treatment interruption among Japanese workers during the COVID-19 pandemic: a cross-sectional study

**DOI:** 10.1101/2021.07.21.21260691

**Authors:** Jun Akashi, Ayako Hino, Seiichiro Tateishi, Tomohisa Nagata, Mayumi Tsuji, Akira Ogami, Shinya Matsuda, Masaharu Kataoka, Yoshihisa Fujino, CORoNaWork Project

**Affiliations:** Second Department of Internal Medicine, School of Medicine, University of Occupational and Environmental Health, Japan; Department of Mental Health, Institute of Industrial Ecological Sciences, University of Occupational and Environmental Health, Japan; Department of Occupational Medicine, School of Medicine, University of Occupational and Environmental Health, Japan; Department of Occupational Health Practice and Management, Institute of Industrial Ecological Sciences, University of Occupational and Environmental Health, Japan; Department of Environmental Health, School of Medicine, University of Occupational and Environmental Health, Japan; Department of Work Systems and Health, Institute of Industrial Ecological Sciences, University of Occupational and Environmental Health, Japan; Department of Preventive Medicine and Community Health, School of Medicine, University of Occupational and Environmental Health, Japan; Department of Environmental Epidemiology, Institute of Industrial Ecological Sciences, University of Occupational and Environmental Health, Japan

**Keywords:** COVID-19, Japan, Patient Acceptance of Health Care, Treatment Refusal, Regional Medical Programs

## Abstract

**Objectives:** This study aimed to examine the relationship between regional infection level and treatment interruption for chronic diseases.

**Methods:** A cross-sectional Internet monitoring survey was performed between December 22 and 26, 2020. Data from 9,510 (5,392 males and 4,118 females) participants needing regular treatment or hospital visits were analyzed. We determined the age-sex- and multivariate-adjusted odds ratios (ORs) of treatment interruption associated with various indices of infection level by nesting multilevel logistic models in prefecture of residence. In the multivariate model, sex, age, marital status, job type, equivalent household income, education, self-rated health, and anxiety were adjusted.

**Results:** The ORs of treatment interruption for the lowest versus highest levels of infection were 1.32 (95% CI: 1.09–1.59) for the overall incidence rate (per 1,000 population), 1.34 (95% CI: 1.10–1.63) for the overall number of people infected, 1.28 (95% CI: 1.06–1.54) for the monthly incidence rate (per 1,000 population), and 1.38 (95% CI: 1.14–1.67) for the number of people infected per month. For each index of infection level, higher infection was linked to more workers experiencing treatment interruption.

**Conclusion:** Higher local infection levels were linked to more workers experiencing treatment interruption. Our results suggest that apart from individual characteristics such as socioeconomic and health status, treatment interruptions during the pandemic were also subject to contextual effects related to regional infection levels. Preventing community spread of COVID-19 may thus protect individuals from indirect effects of the pandemic, such as treatment interruption.

## Introduction

COVID-19, first identified at the end of 2019, is continuing to rage around the world [1–4]. Japan has experienced four waves of the disease through June 2021. In addition to direct effects of severe pneumonia and acute respiratory failure, COVID-19 has also had indirect health effects. COVID-19-related treatment interruption, particularly in patients with chronic diseases, is an emerging issue in several countries [5.6], including Japan [7]. Studies have reported a significant decrease in the number of prescriptions during the pandemic compared to before, and that 40% of patients requiring regular visits have been seen less frequently [8,9].

Treatment interruption can cause serious health care problems in several ways. First, it can worsen the medical condition of patients with chronic diseases that require regular management. Second, fewer opportunities for regular physical examinations may lead to undiagnosed complications and delayed treatment. Further, such medical problems, which could have been avoided by continued treatment, increases medical costs [10]. Studies performed during the COVID-19 pandemic have reported that treatment interruption among patients with chronic diseases is associated with a variety of factors, including fear of becoming infected when seeing the doctor [11,12], scheduling changes by hospitals [13,14], and shortage of medical resources [11]. These factors presumably have differing degrees of impact depending on the level of infection in the region. In addition, patients with unstable socioeconomic status are more likely to discontinue treatment [7,15,16]. Areas with higher prevalence of COVID-19 may be more affected by the loss of job security and other factors that affect individuals with unstable socioeconomic status.

In Japan, the spread of COVID-19 has varied widely by region in terms of the scale of infection and the speed of spread [17.18]. We hypothesize that differences in regional infection rates will affect treatment interruption in each region. The level of infection in a community may directly or indirectly affect fear of visiting medical institutions, anxiety about going out, and financial difficulties, which may cause treatment interruption. For example, the number of people infected with COVID-19 is reported daily by region. Such information will arouse some degree of anxiety and fear in people living in regions with high levels of infection about the safety of the area and the disease. Tokyo, which has recorded the greatest number of infections in Japan, saw a significant drop in prescriptions through May 2020 [8]. Given that pandemics are known to overwhelm medical resources [19], Japan’s lack of capacity to conduct COVID-19 tests in areas with high levels of infection and limited hospital beds has exposed the limits of the country’s medical resources [20].

However, the relationship between regional COVID-19 infection level and treatment interruption remains to be elucidated. Japan provides an ideal opportunity to test our hypotheses due to the country’s large regional variation in COVID-19 infection levels. Therefore, we investigated the relationship between local infection level and treatment interruption in Japan.

## Materials and Methods

### Study Design and Subjects

A cross-sectional study of Internet monitors was conducted from December 22 to 26, 2020, the period corresponding to Japan’s third wave of infection. Data were obtained from participants who indicated they were employed at the time of the survey, with participants selected based prefecture of residence, job type, and sex. A detailed description of the protocol of this survey is provided elsewhere [21]. Of the 33,302 participants in the survey, 6,266 were excluded for providing fraudulent responses. Of the 27,036 remaining participants, data from 9,510 (5392 males and 4118 females) who stated they needed regular treatment or hospital visits were analyzed.

This study was approved by the Ethics Committee of the University of Occupational and Environmental Health, Japan (reference No. R2-079 and R3-006). Participants provided informed consent by completing a form on the survey website.

### Treatment status

We used a single-item question to assess participants’ treatment status: “Do you have a condition that requires regular hospital visits or treatment?” Participants chose from “I do not have such a condition,” “I am continuing with hospital visits and treatment as scheduled,” and “I am not able to continue with hospital visits and treatment as scheduled.”

### Infection level indices

The infection level in each participant’s prefecture of residence was assessed based on the incidence rate for the entire period (per 1,000 population), the number of people infected for the entire period, the incidence rate in one month (per 1,000 population), and the number of infected people in one month.

### Socioeconomic status, health status, and anxiety

Socioeconomic status, health status, and anxiety were assessed through questionnaires in the Internet survey. Socioeconomic factors were age, sex, marital status (married, unmarried, bereaved/divorced), occupation (mainly desk work, mainly interpersonal communication, mainly labor), education, and equivalent income (household income divided by the square root of household size). Health and psychological factors were assessed through participants’ self-rated health status and anxiety about contracting COVID-19. We used the following question to assess anxiety: “Do you feel anxious about being infected with COVID-19?” Participants chose from “yes” or “no.”

### Statistical analysis

We estimated age-sex- and multivariate-adjusted odds ratios (ORs) of treatment interruption associated with regional infection level by nesting multilevel logistic models in prefecture of residence. We used four indices of regional infection level: incidence rate for the entire period (per 1,000 population), number of people infected for the entire period, incidence rate in one month (per 1,000 population), and number of people infected in one month. For analysis, these indices were divided into quartiles and used as area-level variables. In the multivariate model, sex, age, marital status, job type, equivalent household income, education, self-rated health, and anxiety were adjusted. p<0.05 indicated statistical significance. All analyses were conducted using Stata (Stata Statistical Software: Release 16; StataCorp LLC, TX, USA).

## Results

The participants’ characteristics together with residential area according to the number of people infected for the entire period are summarized in Table 1. We stratified the 9,510 participants in need of regular treatment into four groups according to the regional infection level. Socioeconomic factors including sex, age, marital status, household income, education, and occupation, and self-assessment of health status and anxiety related to COVID-19 infection were similar among the four groups.

**Table 1.** Basic characteristics of the study subjects

|  | Residential area according to the number of infected people for the entire period |  |  |  |
| --- | --- | --- | --- | --- |
|  | 74-492 | 507-1496 | 1673-11982 | 12381-52382 |
| Number of subjects | 2130 | 2579 | 2422 | 2379 |
| Age, median (IQR) | 51 (42, 57) | 51 (43, 57) | 52 (44, 58) | 53 (46, 58) |
| Sex, male | 1181 (55.4%) | 1440 (55.8%) | 1403 (57.9%) | 1368 (57.5%) |
| Marital status, married | 1252 (58.8%) | 1496 (58.0%) | 1370 (56.6%) | 1284 (54.0%) |
| Annual equivalent house hold income (JPY) |  |  |  |  |
| 500000-2650000 | 773 (36.3%) | 875 (33.9%) | 816 (33.7%) | 717 (30.1%) |
| 2650000-4500000 | 690 (32.4%) | 824 (32.0%) | 733 (30.3%) | 648 (27.2%) |
| >4500000 | 667 (31.3%) | 880 (34.1%) | 873 (36.0%) | 1014 (42.6%) |
| Education |  |  |  |  |
| Junior high school | 26 (1.2%) | 32 (1.2%) | 30 (1.2%) | 36 (1.5%) |
| High School | 703 (33.0%) | 750 (29.1%) | 619 (25.6%) | 500 (21.0%) |
| Vocational school/college,university,graduate school | 1401 (65.8%) | 1797 (69.7%) | 1773 (73.2%) | 1843 (77.5%) |
| Jobtype |  |  |  |  |
| Mainly desk work | 1144 (53.7%) | 1293 (50.1%) | 1222 (50.5%) | 1264 (53.1%) |
| Jobs mainly involving interpersonal communication | 480 (22.5%) | 590 (22.9%) | 622 (25.7%) | 614 (25.8%) |
| Mainly labor | 506 (23.8%) | 696 (27.0%) | 578 (23.9%) | 501 (21.1%) |
| Self-rated health |  |  |  |  |
| Very good | 742 (34.8%) | 895 (34.7%) | 895 (37.0%) | 885 (37.2%) |
| Neither | 919 (43.1%) | 1104 (42.8%) | 991 (40.9%) | 986 (41.4%) |
| Not good | 469 (22.0%) | 580 (22.5%) | 536 (22.1%) | 508 (21.4%) |
| Do you feel anxious about being infected with COVID-19? |  |  |  |  |
| Yes | 1684 (79.1%) | 2083 (80.8%) | 1904 (78.6%) | 1850 (77.8%) |
| The incidence rate for the entire period (per 1000 of the population), median (IQR) | .28 (.22, .34) | .55 (.51, .59) | 1.26 (.79, 1.51) | 3.12 (1.91, 3.76) |
| The number of infected people for the entire period, median (IQR) | 379 (330, 445) | 1053 (671, 1124) | 2455 (2168, 8438) | 27500 (14427, 52382) |
| The incidence rate for one month (per 1000 of the population), median (IQR) | .09 (.058, .14) | .23 (.16, .32) | .47 (.33, .59) | 1.06 (.74, 1.06) |
| The number of infected people for one month, median (IQR) | 124 (39, 171) | 440 (282, 501) | 1705 (916, 2936) | 9851 (5596, 14690) |

The association between the regional infection level and treatment interruption is summarized in Table 2. According to multivariate analysis, the ORs of treatment interruption for the lowest versus highest regional infection level were 1.32 (95% CI: 1.09–1.59; p=0.003) for the overall incidence rate (per 1,000 population), 1.34 (95% CI: 1.10–1.63; p=0.002) for the overall number of people infected, 1.28 (95% CI: 1.06–1.54; p=0.013) for the monthly incidence rate (per 1,000 population), and 1.38 (95% CI: 1.14–1.67; p=0.001) for the number of people infected per month. For each index of infection level, a higher infection level was linked to more workers experiencing treatment interruption for chronic diseases in Japan. The results remained unchanged after adjusting for age and sex.

**Table 2.** Association between regional COVID-19 infection levels and treatment interruption

|  | Age-sex adjusted |  |  |  | Multivariate* |  |  |  |
| --- | --- | --- | --- | --- | --- | --- | --- | --- |
|  | OR | 95%CI |  | p | OR | 95%CI |  | p |
| The incidence rate for the entire period (per1000) |  |  |  |  |  |  |  |  |
| .10 - .44 | reference |  |  |  | reference |  |  |  |
| .49 - .68 | 1.00 | 0.83 | 1.21 | 0.999 | 1.00 | 0.83 | 1.21 | 0.993 |
| .76 - 1.63 | 1.06 | 0.88 | 1.28 | 0.555 | 1.07 | 0.88 | 1.30 | 0.505 |
| 1.89 - 3.76 | 1.25 | 1.04 | 1.50 | 0.019 | 1.32 | 1.09 | 1.59 | 0.005 |
|  |  |  |  | 0.013† |  |  |  | 0.003† |
| The number of infected people for the entire period |  |  |  |  |  |  |  |  |
| 74 - 492 | reference |  |  |  | reference |  |  |  |
| 507 - 1496 | 1.07 | 0.89 | 1.29 | 0.473 | 1.06 | 0.87 | 1.29 | 0.545 |
| 1673 - 11982 | 1.14 | 0.94 | 1.38 | 0.180 | 1.15 | 0.94 | 1.40 | 0.168 |
| 12381 - 52382 | 1.28 | 1.06 | 1.55 | 0.011 | 1.34 | 1.10 | 1.63 | 0.003 |
|  |  |  |  | 0.008† |  |  |  | 0.002† |
| The incidence rate for one month (per1000) |  |  |  |  |  |  |  |  |
| .018 - .15 | reference |  |  |  | reference |  |  |  |
| .16 - .32 | 1.07 | 0.89 | 1.29 | 0.487 | 1.07 | 0.89 | 1.30 | 0.470 |
| .33 - .61 | 1.05 | 0.87 | 1.27 | 0.641 | 1.06 | 0.87 | 1.29 | 0.576 |
| .66 - 1.12 | 1.21 | 1.02 | 1.45 | 0.032 | 1.28 | 1.06 | 1.54 | 0.009 |
|  |  |  |  | 0.044† |  |  |  | 0.013† |
| The number of infected people for one month |  |  |  |  |  |  |  |  |
| 13 - 203 | reference |  |  |  | reference |  |  |  |
| 204 - 626 | 1.11 | 0.92 | 1.34 | 0.284 | 1.12 | 0.93 | 1.36 | 0.241 |
| 704 - 4373 | 1.16 | 0.96 | 1.40 | 0.127 | 1.18 | 0.97 | 1.43 | 0.093 |
| 5218 - 14690 | 1.30 | 1.08 | 1.57 | 0.006 | 1.38 | 1.14 | 1.67 | 0.001 |
|  |  |  |  | 0.006† |  |  |  | 0.001† |

\* The multivariate model was adjusted for age, sex, marital status, equivalent household income, educational level, jobtype, self-rated health and anxiety about infection
† p for trend

## Discussion

We found that higher regional levels of COVID-19 infection in Japan were correlated with more workers with diseases requiring regular hospital visits and treatment experiencing treatment interruption. To our knowledge, this is the first report to show that community infection levels are associated with treatment interruption.

It is important to emphasize that the association between infection level and treatment interruption remained after adjusting for individual factors such as socioeconomic and health status. These results suggest that apart from individual characteristics, treatment interruptions during the COVID-19 pandemic were also subject to contextual effects related to regional infection levels. For example, while anxiety related to fear of becoming infected during a medical visit is a personal reaction, individuals living in areas with high levels of infection are likely to feel more anxious than those living in areas with low levels of infection. Rescheduling by medical institutions and health care providers is also expected to occur in areas with higher infection levels. Additionally, the COVID-19 pandemic is affecting individuals’ socioeconomic status, which is determined by factors such as employment instability. Higher levels of infection have greater socioeconomic impact, which may be a factor affecting treatment interruption. Our findings are consistent with those of a previous study showing that such individual factors influence treatment interruption [7]. Thus, our study demonstrates that local spread of COVID-19 infection may affect the behavioral characteristics of workers living in the area. These findings suggest that, in addition to an individual patient approach, a population strategy is also needed to prevent the spread of infection and to avoid treatment interruption for manageable diseases.

In this study, both the number of infected people by region and infection rate were associated with treatment interruption. This suggests that it would be informative to report the incidence rate based on the infection status in each region, which reflects the population of that region. However, Japanese news reports tend to emphasize the number of infected people rather than the infection rate by region, the latter of which may contribute to changing the behavior of more people. A previous study reported that Japanese people have greater trust in local information [22], suggesting that reporting the number of infections by region will have a strong influence on individual’s behavioral changes and risk perception.

Increased treatment interruption in areas with high levels of infection may cause further strain on future health care resources. Delaying and avoiding treatment can result in poorer management of chronic diseases, fewer regular checkups, and missed or delayed start of therapy for deteriorating health conditions. It can also lead to increased complications and poor prognosis. These factors in turn can increase future health care needs in the region. The strain on local health care resources due to the COVID-19 pandemic is a serious challenge, and treatment interruption may be an indirect burden on health care resources due to COVID-19. Thus, reducing treatment interruption for manageable diseases may alleviate downstream consequences on the health care system.

The findings of this study indicate that controlling the level of infection in a community has important implications for treatment interruption. With the COVID-19 pandemic expected to continue for some time, sustained control of community-level spread will protect populations from the indirect effects of COVID-19, which include treatment interruption. In addition, strategies are needed to prevent treatment interruption. For example, telemedicine has and will continue to play a major role in the provision of health care during the COVID-19 pandemic [23–26]. Furthermore, educating patients to avoid treatment interruption and widespread use of long-term prescriptions to prevent patients from running out of regular medications will help reduce medical problems caused by treatment interruption.

A major strength of this study was the relatively large sample size, which allowed us to show, for the first time, an association between community infection level and treatment interruption.

However, this study also had several limitations. First, because we conducted a cross-sectional study, causality could not be determined. However, since it is theoretically unlikely that treatment interruption experienced by an individual will increase the COVID-19 infection rate in a region, we think it is likely that high regional infection rates cause treatment interruption. Second, we did not identify workers’ reasons for discontinuing treatment in this study. As discussed above, there are various possible causes of treatment interruption, which may vary by region. Third, we did not inquire about the diseases being treated. Treatment interruption may vary depending on the presence or absence of symptoms and the potential disadvantages of discontinuing treatment for a particular disease.

## Conclusion

We found that higher regional infection levels were linked to more workers experiencing treatment interruption during the third wave of COVID-19 infection in Japan. Our findings suggest that in addition to individual factors such as socioeconomic status and health status, high regional infection levels may contribute to behavioral changes in the local population, leading to treatment interruption. Preventing community spread of COVID-19 may thus be useful for avoiding treatment interruption for chronic diseases, an emerging medical problem brought about by COVID-19.

## Data Availability

Data not available due to ethical restrictions.

## Data statement

Data not available due to ethical restrictions.

## Acknowledgements

The current members of the CORoNaWork Project, in alphabetical order, are as follows: Dr. Yoshihisa Fujino (present chairperson of the study group), Dr. Akira Ogami, Dr. Arisa Harada, Dr. Ayako Hino, Dr. Hajime Ando, Dr. Hisashi Eguchi, Dr. Kazunori Ikegami, Dr. Kei Tokutsu, Dr. Keiji Muramatsu, Dr. Koji Mori, Dr. Kosuke Mafune, Dr. Kyoko Kitagawa, Dr. Masako Nagata, Dr. Mayumi Tsuji, Ms. Ning Liu, Dr. Rie Tanaka, Dr. Ryutaro Matsugaki, Dr. Seiichiro Tateishi, Dr. Shinya Matsuda, Dr. Tomohiro Ishimaru, and Dr. Tomohisa Nagata. All members are affiliated with the University of Occupational and Environmental Health, Japan.

## References

1. Guan WJ, Ni ZY, Hu Y, Liang WH, Ou CQ, He JX, et al. Clinical Characteristics of Coronavirus Disease 2019 in China. N Engl J Med. 2020;382(18):1708–20.

2. Holshue ML, DeBolt C, Lindquist S, Lofy KH, Wiesman J, Bruce H, et al. First Case of 2019 Novel Coronavirus in the United States. N Engl J Med. 2020;382(10):929–36.

3. Phan LT, Nguyen TV, Luong QC, Nguyen TV, Nguyen HT, L. HQ, et al. Importation and Human-to-Human Transmission of a Novel Coronavirus in Vietnam. N Engl J Med. 2020;382(9):872–4.

4. Rothe C, Schunk M, Sothmann P, Bretzel G, Froeschl G, Wallrauch C, et al. Transmission of 2019-nCoV Infection from an Asymptomatic Contact in Germany. N Engl J Med. 2020;382(10):970–1.

5. Birkmeyer JD, Barnato A, Birkmeyer N, Bessler R, Skinner J. The Impact Of The COVID-19 Pandemic On Hospital Admissions In The United States. Health Aff. 2020;39(11):2010–2017.

6. Czeisler M, Marynak K, Clarke KEN, Salah Z, Shakya I, Thierry JM, et al. Delay or Avoidance of Medical Care Because of COVID-19-Related Concerns - United States, June 2020. MMWR Morb Mortal Wkly Rep. 2020;69(36):1250–7.

7. Fujimoto K, Ishimaru T, Tateishi S, Nagata T, Tsuji M, Eguchi H, et al. A cross-sectional study of socioeconomic status and treatment interruption among Japanese workers during the COVID-19 pandemic. medRxiv. 2021:2021.02.22.21252190.

8. LoPresti M, Seo T, Sato N. Pandemics and Access to Care: Use of Real-World DATA to Examine the IMPACT of COVID-19 on Pharmacy Visits in JAPAN. Value Health. 2020;23:S685.

9. Takakubo T, Odagiri Y, Machida M, Takamiya T, Fukushima N, Kikuchi H, et al. Changes in the medical treatment status of Japanese outpatients during the coronavirus disease 2019 pandemic. Journal of General and Family Medicine. 2021;00:1–16.

10. Erol MK, Kayıkçıoğlu M, Kılıçkap M, Güler A, Yıldırım A, Kahraman F, et al. Treatment delays and in-hospital outcomes in acute myocardial infarction during the COVID-19 pandemic: A nationwide study. Anatol J Cardiol. 2020;24(5):334–42.

11. Czeisler M, Marynak K, Clarke KEN, Salah Z, Shakya I, Thierry JM, et al. Delay or Avoidance of Medical Care Because of COVID-19-Related Concerns - United States, June 2020. MMWR Morb Mortal Wkly Rep. 2020;69(36):1250–7.

12. Chudasama YV, Gillies CL, Zaccardi F, Coles B, Davies MJ, Seidu S, et al. Impact of COVID-19 on routine care for chronic diseases: A global survey of views from healthcare professionals. Diabetes Metab Syndr. 2020;14(5):965–7.

13. Chopra V, Toner E, Waldhorn R, Washer L. How Should U.S. Hospitals Prepare for Coronavirus Disease 2019 (COVID-19)? Ann Intern Med. 2020;172(9):621–2.

14. Khullar D, Bond AM, Schpero WL. COVID-19 and the Financial Health of US Hospitals. JAMA. 2020;323(21):2127–2128.

15. Feinstein JS. The relationship between socioeconomic status and health: a review of the literature. Milbank Q. 1993;71(2):279–322.

16. Swain GR. How does economic and social disadvantage affect health. Focus. 2016;33(1):1–6.

17. Ministry of Health, Labour and Welfare. Current status of the novel coronavirus infection and the response of the MHLW. 31 December 2020. Available at: https://www.mhlw.go.jp/stf/newpage_15828.html. Accessed June 27, 2021.

18. Furuse Y, Ko YK, Saito M, Shobugawa Y, Jindai K, Saito T, et al. Epidemiology of COVID-19 Outbreak in Japan, from January-March 2020. Jpn J Infect Dis. 2020;73(5):391–3.

19. Emanuel EJ, Persad G, Upshur R, Thome B, Parker M, Glickman A, et al. Fair Allocation of Scarce Medical Resources in the Time of Covid-19. N Engl J Med. 2020;382(21):2049–55.

20. Watanabe M. The COVID-19 Pandemic in Japan. Surg Today. 2020;50(8):787–93.

21. Fujino Y, Ishimaru T, Eguchi H, et al. Protocol for a nationwide Internet-based health survey in workers during the COVID-19 pandemic in 2020. medRxiv. Published online February 5, 2021:2021.02.02.21249309.

22. Muto K, Yamamoto I, Nagasu M, Tanaka M, Wada K. Japanese citizens’ behavioral changes and preparedness against COVID-19: An online survey during the early phase of the pandemic. PLoS One. 2020;15(6):e0234292.

23. Grabowski DC, O’Malley AJ. Use of telemedicine can reduce hospitalizations of nursing home residents and generate savings for medicare. Health Aff (Millwood). 2014;33(2):244–50.

24. Bokolo Anthony J. Use of Telemedicine and Virtual Care for Remote Treatment in Response to COVID-19 Pandemic. J Med Syst. 2020;44(7):132.

25. Neubeck L, Hansen T, Jaarsma T, Klompstra L, Gallagher R. Delivering healthcare remotely to cardiovascular patients during COVID-19: A rapid review of the evidence. Eur J Cardiovasc Nurs. 2020;19(6):486–94.

26. Elson EC, Oermann C, Duehlmeyer S, Bledsoe S. Use of telemedicine to provide clinical pharmacy services during the SARS-CoV-2 pandemic. Am J Health Syst Pharm. 2020;77(13):1005–6.

